# Rotating Night Shift Work and Risk of Incident Cardiovascular Disease and Mortality by Chronotype: A Prospective Cohort Study in Nurses’ Health Study II

**DOI:** 10.64898/2026.09.16.26363233

**Authors:** Sina Kianersi, Yue Liu, Tamar Sofer, Isha Agarwal, Frank A.J.L. Scheer, Marta Guasch-Ferré, Susan Redline, Eva Schernhammer, Kathryn M. Rexrode, Tianyi Huang

## Abstract

**Introduction:** Rotating night shift work is associated with chronic circadian disruption and may contribute to higher cardiovascular disease (CVD) risk and mortality. The impact of rotating night shift work may vary by chronotype, particularly when an individual’s circadian preference is misaligned with work schedules.

**Methods:** In 2009, 69,458 women (mean age 54.4 years) free of CVD in the Nurses’ Health Study II reported chronotype. Duration of rotating night shift work has been assessed biennially since 1989. Participants were followed from 2009 through June 2021 for incident CVD (myocardial infarction or stroke) and all-cause mortality. We used time-varying Cox proportional hazards models to estimate hazard ratios (HRs) and 95% confidence intervals (CIs) for outcomes in relation to rotating night shift work and chronotype and to evaluate whether the associations differed by chronotype. Primary models adjusted for key demographics, CVD risk factors, and workplace factors.

**Results:** We documented 1,005 incident CVD events over 800,630 person-years and 1,777 deaths over 806,043 person-years. Each 5-year increase in rotating night shift work was associated with a modestly higher risk of incident CVD (HR 1.07; 95% CI 1.01, 1.14) and a 13% higher risk of all-cause mortality (HR 1.13; 95% CI 1.08, 1.17). Greater eveningness was associated with higher incident CVD risk (HR 1.17; 95% CI 1.06, 1.28) and all-cause mortality (HR 1.18; 95% CI 1.10, 1.26). For incident CVD, the increased risk per 5-year increment in rotating night shift work was observed only among definite morning types (HR 1.11; 95% CI 0.99, 1.23), but not among definite evening types (HR 0.99; 95% CI 0.86, 1.15; p-interaction=0.1732). For all-cause mortality, the increased risk associated with shorter-term rotating night shift work (<5 years) was more evident among definite morning types (HR 1.39; 95% CI 1.12, 1.73) than definite evening types (HR 1.37; 95% CI 0.98, 1.91). However, this difference was not observed for longer duration of shift work (≥5 years), and the overall interaction was not statistically significant (p-interaction=0.57). Similar patterns were observed when shift work duration was modeled continuously using restricted cubic spline analyses.

**Conclusion:** Rotating night shift work was associated with a modest increase in CVD and all-cause mortality risk. The overall pattern of associations across chronotypes suggested that nurses with a morning chronotype may be more susceptible to the adverse effects of rotating night shift work.

## INTRODUCTION

Non-daytime work schedules are prevalent worldwide, with around 24 million workers engaged in shift work in the United States during 2017-2018 (1). Systematic and clinical reviews have recognized night shift work as a risk factor for several adverse health outcomes, including diabetes, cardiovascular diseases (CVD), and all-cause mortality, though findings from individual observational studies have not always been consistent (2–6). A key mechanism underlying these observations is chronic recurrent circadian misalignment, a condition where the internal circadian body clock is out of sync with the external physical (e.g., light) and behavioral (e.g., work schedules) cycles (7–10). In turn, circadian misalignment disrupts hormone secretion, metabolic regulation, cardiovascular functions and health behaviors, and over the long term, culminates in a spectrum of cardiometabolic risk factors and increased mortality (11–15).

Chronotype refers to one’s propensity of earlier or later sleeping times and partially reflects the timing of the internal body clock (i.e., circadian timing system) while also being shaped by behavioral, social, and environmental factors (16, 17). In previous work, we observed that an evening chronotype, compared with morning or intermediate chronotype, was associated with worse cardiometabolic profiles, including both risk behaviors and clinical disorders (18, 19). A mismatch between chronotype and work schedules occurs when work hours interfere with an individual’s preferred sleeping times (20). This mismatch may manifest among both night shift workers, especially those with a morning chronotype and daytime workers (particularly those having early morning shifts) with an evening chronotype, potentially exacerbating the underlying circadian misalignment (21). Therefore, simultaneous consideration of night shift work and chronotype is essential to more accurately characterize the health impact of circadian misalignment (22). However, limited studies have evaluated the mismatch between chronotype and work schedule in relation to health outcomes. Prior studies have reported associations of work schedule-chronotype mismatch with adverse sleep, metabolic, and cardiometabolic factors (20, 21, 23), although findings have not always been consistent; however, prospective evidence in large cohorts, especially for CVD and all-cause mortality, remains limited.

In the current study, we prospectively examined the independent associations of rotating night shift work history and chronotype with incident CVD (myocardial infarction and stroke in this study) and all-cause mortality and their interactions in the Nurses’ Health Study II (NHSII), a large cohort study with repeated assessments of work schedules, chronotype, and behavioral and clinical factors. We hypothesized that (1) a history of rotating night shift work (operationalized using multiple metrics such as duration, current rotating night shift work status, frequency, and the less commonly examined metric of time since quitting rotating night shift work) is associated with higher risks of CVD and all-cause mortality in a dose-response manner; (2) evening chronotype is associated with higher CVD and mortality risk compared with intermediate chronotype; and (3) CVD and all-cause mortality risks are greatest when chronotype is mismatched with the work schedule, with stronger associations of rotating night shift work with CVD and all-cause mortality among participants with a morning chronotype than among those with intermediate or evening chronotypes.

## METHODS

### Study Design, Setting, and Participants

We conducted a prospective cohort study from 2009 to 2021 in NHSII. Established in 1989, this cohort began with the enrollment of 116,429 primarily white (>90%) female registered nurses aged 25–42 years from 14 U.S. states (24). Participants completed lifestyle and health questionnaires at baseline and every 2 years thereafter, with invitations and data collection conducted mainly by mail and supplemented by telephone and web-based formats; follow-up completion rates exceeded 85% across questionnaire cycles (25). We excluded 25,972 participants who died before 2009, withdrew from the study, or did not return the 2009 questionnaire, which served as baseline for the present study. We also excluded 2,285 participants with no follow-up data after 2009. We further excluded 7,047 participants with prevalent cancer (excluding basal cell carcinoma and squamous cell carcinoma), 775 with a history of stroke and/or myocardial infarction (MI) at baseline, and 10,892 with missing chronotype data, leaving 69,458 participants for the current analysis (Supplemental Figure 1). The study protocol was approved by the institutional review boards of the Brigham and Women’s Hospital and Harvard T.H. Chan School of Public Health, and those of participating registries as required. We reported this study in accordance with STROBE guidelines (26).

### Shift work

We created multiple metrics for characterizing rotating night shift work exposure as used in previous studies (5, 18, 20, 27). The primary exposure was lifetime duration of rotating night shift work. In 1989, participants reported the total number of years they had ever worked rotating night shifts: “*What is the total number of years during which you worked rotating night shifts (at least 3 nights/month in addition to days or evenings in that month)?*”. In subsequent questionnaires (1991, 1993, 1997, 2001, 2005, 2007, 2011, 2013), participants reported the number of months worked on rotating night shifts since the previous questionnaire; “*Since [previous questionnaire date], how many months have you worked ROTATING night shifts (at least 3 nights/month in addition to other days and evenings in that month)?*” Possible responses included six options (None, “1-4 months”, “5-9”, “10-14”, “15-19”, “20+”). We assigned the midpoint to represent each response category (e.g., assigning 7 months for “5–9 months”, and 20 months for “20+”) and calculated cumulative years of rotating night work by summing these months across all prior questionnaires and dividing it by 12. If shift work response in a given questionnaire cycle was missing, we carried forward the value from the immediately preceding cycle once. We then categorized cumulative years of rotating night work at each follow up cycle as none, <5 years, or ≥5 years for the primary analyses.

Using the same cumulative months measure, we defined a time varying indicator of ever having worked rotating night shifts (ever vs never) and additionally created a four level duration variable (none, <5, ≥5 to <10, ≥10 years of rotating night shift work) to capture the impact of long-term exposure to shift work. Following a prior NHSII study (5), we derived a time varying measure of time since quitting rotating night shift work, defined as the years elapsed since the last reported rotating night shift episode among former workers and categorized as never worked rotating nights, currently working rotating nights, <12 years, 12–24 years, or >24 years since quitting rotating night shift work. We also collapsed this measure into a three level shift work status variable (never, current, or past rotating night shift work). In 2009, participants also reported their usual night shift frequency in their current job (“*On average, how many night shifts did you work per month?*”, defined as most work hours between 11pm and 7am). We assigned midpoints to the response categories to derive a continuous nights/month measure and additionally categorized this baseline frequency as none, 1–6, or ≥7 nights/month (cut at the median, corresponding to the 5–6 nights/month category). Finally, to assess robustness of imputing missing shift work data, we also constructed an alternative duration measure restricted to women with non missing shift work information across all questionnaires.

### Chronotype

Chronotype was assessed, as a time-dependent variable, in 2009 and 2015 using one representative question from the Horne–Östberg Morningness–Eveningness Questionnaire (MEQ) (28). This single-item self-classification correlates strongly with the overall MEQ score (r > 0.7) (29, 30): “*One hears about morning and evening types of people. Which ONE of these types do you consider yourself to be?*” This question had five response options (Definitely a morning type, More of a morning than an evening type, More of an evening than a morning type, Definitely an evening type, Neither). We combined the categories following previously published NHSII papers (18, 20), and recoded the responses to three groups, definite morning, intermediate (more evening, neither, and more morning), and definite evening.

### Incident Cardiovascular Diseases

Incident CVD was defined as the first confirmed fatal or nonfatal MI or stroke after return of the 2009 questionnaire. Potential MI and stroke events were identified through biennial self-reports and deaths reported by families or the postal service and/or identified through the National Death Index. For reported events, we requested permission to obtain medical records; for deaths, we obtained death certificates and sought supporting medical or autopsy records when available. When complete documentation was unavailable, participant reports, including self-reports and/or participant-corroborated diagnoses based on additional details obtained by letter/interview, were used. Study physicians blinded to participant characteristics reviewed available documentation to confirm MI according to World Health Organization criteria: typical symptoms plus diagnostic ECG changes and/or elevated cardiac biomarkers, including troponin when accompanied by symptoms or ECG changes (31, 32). Strokes were confirmed and classified, when documentation permitted, according to National Survey of Stroke criteria: a sudden or rapid-onset neurologic deficit persisting >24 hours or until death, excluding events due to infection, trauma, malignancy, or radiology-only ‘silent’ strokes (33). The CVD event date was defined as the earliest diagnosis date of MI or stroke.

### All-Cause Mortality

The primary outcome of interest was death from any cause, identified through reports from next-of-kin, notifications from the U.S. Postal Service when mail was returned, and periodic linkage with the National Death Index. For reported deaths, additional information was sought from next-of-kin, medical records, state tumor registries, and death certificates, and investigators who were blinded to shift work exposure, chronotype and other risk factors reviewed all available data to classify the underlying cause of death according to International Classification of Diseases (ICD) codes. Mortality follow-up, including use of the National Death Index, has been previously validated, showing that this approach could reliably ascertain 98% of the deaths in the cohort (34, 35).

### Covariates

The following covariates were included in analyses: age; ethnic background; socioeconomic status (census-tract median family income) (36); marital status; workplace (ICU/ER/OR, in hospital, outpatient, retired/other); menopausal status; postmenopausal hormone use (current, past, never user); family history of MI and/or stroke; aspirin use; smoking status; self-reported sleep duration (<7, 7-8, >8 hours); cumulative average physical activity, measured with a validated questionnaire and quantified as total metabolic equivalent of task (MET) hours per week (37); BMI (kg/m^2^); cumulative average Alternative Healthy Eating Index-2010 without the alcohol component (AHEI-2010) assessed with a 152-item food-frequency questionnaire (38, 39); cumulative average alcohol intake (g/day); and self-reported physician-diagnosed metabolic disorders, including hypertension, type 2 diabetes, and hypercholesterolemia.

### Statistical Analysis

In descriptive analyses, we summarized baseline participant characteristics in the overall cohort and across rotating night shift work duration categories. We used Cox proportional hazards regression to estimate hazard ratios (HRs) and 95% confidence intervals (CIs) for associations of cumulative duration of rotating night shift work and chronotype with incident CVD and all-cause mortality separately. For the CVD analysis, person-time accrued from the return date of the 2009 questionnaire until the date of CVD diagnosis, death, or the end of follow-up (June 2021), whichever occurred first. For the all-cause mortality analysis, person-time was calculated from 2009 questionnaire return date until death or the end of follow-up.

We fit an age-adjusted model and two multivariable models. The age-adjusted model included age as the underlying time scale (in months). Model 1 (the primary model) adjusted for demographic factors, including age, ethnic background, census-tract median family income, marital status, workplace, family history of MI and/or stroke, menopausal status, and postmenopausal hormone therapy use. Model 2 further adjusted for traditional CVD risk factors, including diabetes, hypertension, hypercholesterolemia, aspirin use, smoking status, sleep duration, physical activity, BMI, AHEI-2010, and alcohol use. When available, covariates were treated as time-varying and modeled in their continuous form to reduce measurement error and residual confounding (40), see Supplemental Figure 2. We addressed missing covariate values by carrying forward information from prior questionnaire cycles when available. We evaluated the proportional hazards assumption by likelihood ratio tests comparing nested Cox models with and without time-dependent interaction term between duration of rotating night shift work or chronotype and follow-up time; there was no evidence of violation of the assumption (p-interaction > 0.17).

We implemented Cox models using the SAS MPHREG9 macro, which accommodates time-varying exposures/covariates across questionnaire cycles with ties handled using the Efron method (41). Tests for linear trend were conducted by modeling duration of rotating night shift work (in years) and chronotype (ranging from 1, definite morning, to 3, definite evening) as continuous variables. To assess potential nonlinearity in the association of cumulative rotating night shift work duration (years) with incident CVD and all-cause mortality, we fit restricted cubic splines in Cox models using the SAS %LGTPHCURV9 macro (42), with 3 knots placed at the 5^th^, 50^th^, and 95^th^ percentiles of the exposure distribution.

To evaluate whether associations between cumulative rotating night shift work duration and outcomes differed by chronotype, we conducted stratified analyses by chronotype. We assessed multiplicative interaction by adding a rotating night shift work × chronotype product term and estimating P for interaction using a likelihood ratio test comparing models with versus without the product term. We followed developed guidelines when reporting our interaction analysis findings (43). We also conducted the restricted cubic splines analyses described above in each chronotype category separately.

We conducted the following prespecified secondary analyses to evaluate robustness of the primary findings. First, we repeated the primary analyses using alternative shift work exposure definitions as described above to assess sensitivity to how rotating night shift work was operationalized. Second, we evaluated the associations separately for MI and stroke to evaluate whether associations differed across major CVD endpoints. Third, we evaluated the joint associations using a combined chronotype–shift work variable (reference: no rotating night shift work with intermediate chronotype). All data processing and statistical analyses were conducted in SAS; figures were generated in Python. Statistical tests were two-sided, with α = 0.05.

## RESULTS

### Baseline Characteristics (2009)

Of 69,458 participants, the mean (SD) age was 54.4 (4.6) years, and 97% identified as White (Table 1). Chronotype distribution was 12% definite evening, 35% definite morning, and 54% intermediate. Mean duration of rotating night shift work was 3.5 years (SD: 4.5; range: 0– 36.7 years). Approximately 27% of participants reported no rotating night-shift work; 49% reported <5 years, and 23% reported ≥5 years. Compared with participants reporting no shift work, those with ≥5 years were more likely to report a definite evening chronotype, have lower census-tract median family income, and work in acute-care settings. In addition, longer duration of rotating night shift work was associated with higher BMI and higher prevalence of smoking, suboptimal sleep duration (<7 or >8 h/day), hypertension, and type 2 diabetes.

**Table 1.** Characteristics of participants at baseline by duration of rotating night shift work in the Nurses’ Health Study II (2009)

|  |  |  |  |  |
| --- | --- | --- | --- | --- |
|  | Duration of rotating night shift work (years) |  |  |  |
|  | Total sample<br>N = 69,458 | None<br>N = 19,053 | <5 years<br>N = 34,343 | ≥5 years<br>N = 16,062 |
| **Chronotype** |  |  |  |  |
| - Definite morning, % | 34.7 | 36.3 | 35.0 | 32.2 |
| - Intermediate, % | 53.7 | 54.2 | 53.7 | 53.0 |
| - Definite evening, % | 11.6 | 9.5 | 11.4 | 14.8 |
| **Age (years), Mean (SD)** | 54.4 (4.6) | 54.8 (4.6) | 54.3 (4.7) | 54.2 (4.6) |
| **White, %** | 96.5 | 97.3 | 96.5 | 95.6 |
| **Census-tract median family income (USD), Mean (SD)** | 82693 (32109) | 83926 (32574) | 83718 (32761) | 79041 (29793) |
| **Married or domestic partner, %** | 91.6 | 93.4 | 91.9 | 88.9 |
| **Workplace** |  |  |  |  |
| - ICU/ER/OR, % | 9.7 | 6.9 | 8.8 | 15.2 |
| - In hospital, % | 19.5 | 15.9 | 18.9 | 25.1 |
| - Outpatient, % | 30.8 | 34.1 | 31.6 | 24.9 |
| - Retired/other, % | 40.0 | 43.1 | 40.7 | 34.8 |
| **Family history of MI and/or stroke, %** | 52.6 | 51.9 | 52.0 | 55.0 |
| **Type 2 diabetes, %** | 5.5 | 4.6 | 5.3 | 7.1 |
| **Hypertension, %** | 34.1 | 32.7 | 33.5 | 37.0 |
| **Hypercholesterolemia, %** | 49.9 | 49.9 | 49.4 | 50.8 |
| **Postmenopausal, %** | 67.9 | 69.9 | 66.9 | 67.8 |
| **Postmenopausal hormone therapy** |  |  |  |  |
| - Current user, % | 15.1 | 16.6 | 15.0 | 13.5 |
| - Past user, % | 20.7 | 22.1 | 20.2 | 20.3 |
| - Never user, % | 64.2 | 61.4 | 64.8 | 66.3 |
| **Aspirin use, %** | 33.0 | 31.9 | 32.5 | 35.1 |
| **Currently smoker, %** | 6.0 | 4.8 | 5.9 | 7.6 |
| **Sleep duration <7 or >8 (hours/day), %** | 32.5 | 28.1 | 32.3 | 37.8 |
| **Physical activity (MET-h/week), Mean (SD)** | 20.5 (18.3) | 19.5 (17.5) | 20.6 (18.1) | 21.4 (19.3) |
| **Body Mass Index (kg/m<sup>2</sup>), Mean (SD)** | 27.6 (6.4) | 27 (6) | 27.5 (6.3) | 28.8 (6.9) |
| **Alternate Healthy Eating Index-2010 score, Mean (SD)** | 49.3 (9.3) | 49.5 (9.3) | 49.4 (9.3) | 48.8 (9.2) |
| **Alcohol use (g/day), Mean (SD)** | 4.5 (6.7) | 4.6 (6.9) | 4.6 (6.8) | 4.2 (6.4) |
| ED = emergency department; ICU = intensive care unit; OR = operating room.<br>Physical activity, AHEI-2010 score, and alcohol use are presented as cumulative averages. At baseline, physical activity was averaged across available assessments from 1989, 1991, 1997, 2001, 2005, and 2009; AHEI-2010 score and alcohol intake were represented by averaging the cumulative means from repeated food-frequency questionnaires through 2007 and through 2011. These and other time-varying covariates were updated in subsequent questionnaire cycles. |  |  |  |  |

### Chronotype, Rotating Night Shift Work, and CVD Associations

During 800,630 person-years of follow-up (median: 11.8 years), 1,005 incident CVD cases were documented (Table 2). In age-adjusted analyses, both longer duration of rotating night shift work and greater eveningness were significantly associated with higher CVD risk, although these associations were attenuated after further multivariable adjustment. In Model 1, compared with participants reporting no rotating night shift work, the HRs were 0.98 (95% CI: 0.84, 1.14) for <5 years and 1.21 (95% CI: 1.02, 1.44) for ≥5 years of rotating night shift work. When modeled continuously, each 5-year increase in rotating night shift work duration was associated with a 7% higher CVD risk (95% CI: 1.01, 1.14; p-trend: 0.0208). After further adjustment for cardiometabolic comorbidities and lifestyle factors in Model 2, the association was attenuated (HR per 5-year increase: 1.02; 95% CI: 0.96, 1.09; p-trend: 0.4353). We found no evidence of nonlinearity using the restricted cubic spline model (Figure 1A; p-nonlinearity = 0.99).

**Figure 1.**
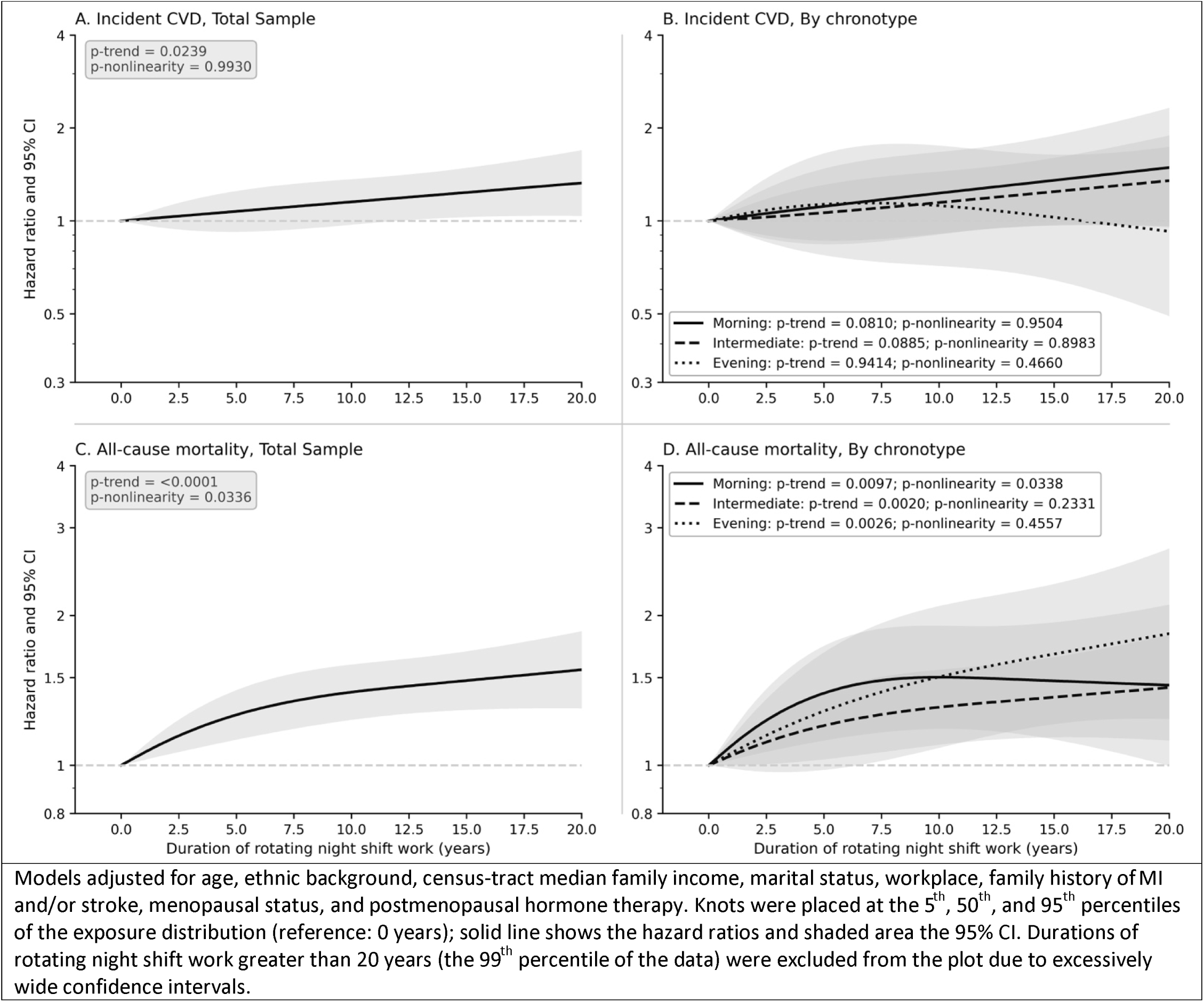
Restricted cubic spline for duration of rotating night shift work in relation to incident cardiovascular disease and all-cause mortality, N = 69,458.

**Table 2.**
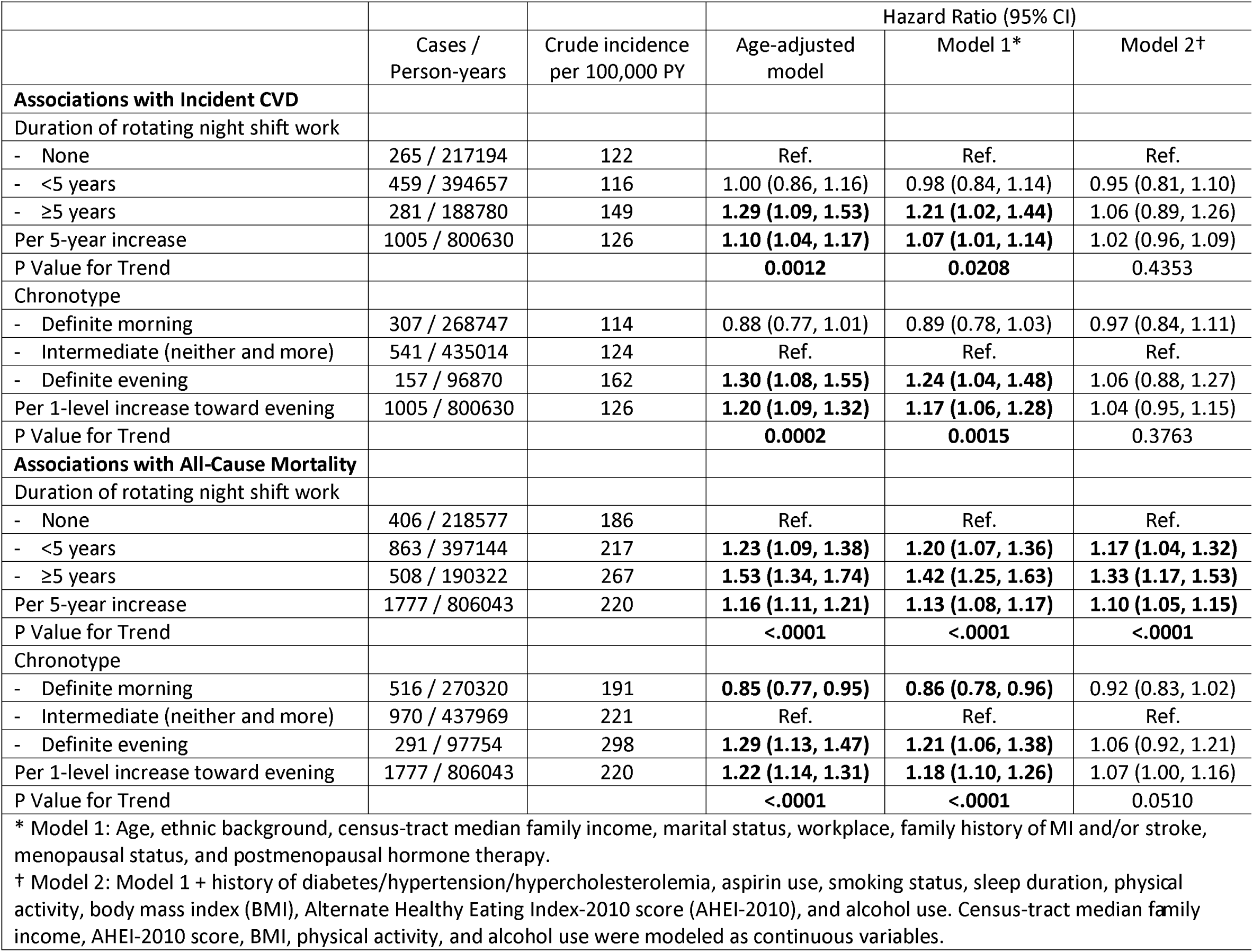
Prospective associations between rotating night shift work, chronotype, and risks of cardiovascular disease and all-cause mortality in the NHSII (N = 69,458)

Compared with participants reporting an “intermediate” chronotype, the HRs were 0.89 (95% CI: 0.78, 1.03) for “definite morning” and 1.24 (95% CI: 1.04, 1.48) for “definite evening” chronotypes in Model 1. When modeled continuously, each 1-level increase toward eveningness was associated with 17% higher CVD risk (95% CI: 1.06, 1.28; p-trend: 0.0015). After additional adjustment for lifestyle factors and comorbidities in Model 2, the association was attenuated and no longer statistically significant (HR per 1-level increase: 1.04; 95% CI: 0.95, 1.15; p-trend: 0.3763).

No evidence of significant multiplicative interaction between chronotype and duration of rotating night shift work was observed for incident CVD (P=0.1732; Table 3). However, we only observed a suggestion of positive associations between shift work duration and CVD risk among definite morning chronotypes (HR per 5-year increase in rotating night shift work: 1.11; 95% CI: 0.99, 1.23) and, to some extent, intermediate chronotypes (HR: 1.08; 95% CI: 0.99, 1.17), but no association among definite evening chronotypes (HR: 0.99; 95% CI: 0.86, 1.15). Similar patterns were observed in the restricted cubic spline analyses (Figure 1B).

**Table 3.** Associations of rotating night shift work with incident CVD and all-cause mortality, stratified by chronotype (NHSII; N = 69,458)

|  |  |  |  |  |  |  |
| --- | --- | --- | --- | --- | --- | --- |
|  | Sub-group analysis by chronotype |  |  |  |  |  |
| Incident CVD | Definite morning |  | Intermediate |  | Definite evening |  |
| Duration of rotating night shift work | Cases | HR (95% CI) | Cases | HR (95% CI) | Cases | HR (95% CI) |
| - None | 90 | Ref. | 143 | Ref. | 32 | Ref. |
| - <5 years | 133 | 0.87 (0.66, 1.14) | 247 | 0.99 (0.81, 1.22) | 79 | 1.31 (0.86, 1.99) |
| - ≥5 years | 84 | 1.25 (0.92, 1.69) | 151 | 1.23 (0.97, 1.55) | 46 | 1.22 (0.76, 1.95) |
| Per 5-year increase | 307 | 1.11 (0.99, 1.23) | 541 | 1.08 (0.99, 1.17) | 157 | 0.99 (0.86, 1.15) |
| p for interaction | 0.17 |  |  |  |  |  |
| All-Cause Mortality |  |  |  |  |  |  |
| Duration of rotating night shift work | Cases | HR (95% CI) | Cases | HR (95% CI) | Cases | HR (95% CI) |
| - None | 116 | Ref. | 240 | Ref. | 50 | Ref. |
| - <5 years | 272 | **1.39 (1.12, 1.73)** | 457 | 1.08 (0.92, 1.26) | 134 | 1.37 (0.98, 1.91) |
| - ≥5 years | 128 | **1.49 (1.15, 1.93)** | 273 | **1.29 (1.08, 1.55)** | 107 | **1.72 (1.21, 2.44)** |
| Per 5-year increase | 516 | **1.12 (1.03, 1.22)** | 970 | **1.10 (1.04, 1.17)** | 291 | **1.17 (1.06, 1.28)** |
| p for interaction | 0.57 |  |  |  |  |  |
| Models adjusted for age, ethnic background, census-tract median family income, marital status, workplace, family history of MI and/or stroke, menopausal status, and postmenopausal hormone therapy. |  |  |  |  |  |  |
| P for multiplicative interaction between chronotype and duration of rotating night shift work was estimated using a likelihood ratio test comparing models with and without the chronotypexshift work product term (both modeled as continuous variables); HR: Hazard Ratio; CVD: Cardiovascular Disease |  |  |  |  |  |  |

### Chronotype, Rotating Night Shift Work, and All-Cause Mortality

During 806,043 person-years of follow-up (median: 11.8 years), 1,777 deaths were documented (Table 2). Both longer duration of rotating night shift work and greater eveningness were significantly associated with all-cause mortality in age-adjusted and Model 1 analyses. In Model 1, compared with participants reporting no rotating night shift work, the HRs were 1.20 (95% CI: 1.07, 1.36) for <5 years and 1.42 (95% CI: 1.25, 1.63) for ≥5 years of rotating night shift work. When modeled continuously, each 5-year increase in rotating night shift work duration was associated with 13% higher all-cause mortality (95% CI: 1.08, 1.17; p-trend <0.0001). After further adjustment for cardiometabolic comorbidities and lifestyle factors in Model 2, the association was attenuated but remained statistically significant (HR per 5-year increase: 1.10; 95% CI: 1.05, 1.15; p-trend <0.0001). The restricted cubic spline analysis showed a strong positive association that was approximately linear, with weak evidence of nonlinearity (Figure 1C; p-trend <0.0001; p-nonlinearity = 0.034).

In Model 1, compared with those with an “intermediate” chronotype, the HRs for all-cause mortality were 0.86 (95% CI: 0.78, 0.96) for “definite morning” and 1.21 (95% CI: 1.06, 1.38) for “definite evening” chronotype. Each 1-level increase toward eveningness was associated with 18% higher mortality risk (95% CI: 1.10, 1.26; p-trend <0.0001). After additional adjustment for lifestyle and clinical factors in Model 2, the association was attenuated and borderline nonsignificant (HR per 1-level increase: 1.07; 95% CI: 1.00, 1.16; p-trend: 0.0510).

No evidence of multiplicative interaction between chronotype and duration of rotating night shift work was observed for all-cause mortality (P=0.57). In stratified analyses, rotating night shift work was associated with higher all-cause mortality within every chronotype group (Table 3). Estimates per 5-year increase in shift work duration were comparable across chronotype groups: definite morning (HR: 1.12; 95% CI: 1.03, 1.22), intermediate (HR: 1.10; 95% CI: 1.04, 1.17), and definite evening (HR: 1.17; 95% CI: 1.06, 1.28). However, in categorical analyses, the association for <5 years of rotating night shift work (vs none) was stronger and more precise among definite morning types (HR: 1.39; 95% CI: 1.12, 1.73) than definite evening types (HR: 1.37; 95% CI: 0.98, 1.91). This pattern was supported by restricted cubic spline analyses stratified by chronotype (Figure 1D): among morning chronotypes, mortality risk increased more sharply with shorter duration of shift work but tended to plateau with longer durations, whereas among intermediate or evening chronotypes, mortality risk increased more linearly with shift work duration.

### Additional Analyses

Similar results were found when we used alternative definitions for shift work (Supplemental Tables 1 and 2). Additionally, consistent results were found when we evaluated MI and stroke outcomes separately (Supplemental Tables 3 and 4). In joint analyses, participants with definite morning chronotype and <5 years of rotating night shift work had the lowest risk of incident CVD, while those with definite morning chronotype and no rotating night shift work had the lowest risk of all-cause mortality (Supplemental Table 5). Finally, findings were similar when we restricted our analysis to participants with complete (no missing values) rotating night shift work information (Supplemental Tables 1 and 2).

## DISCUSSION

In this prospective analysis of NHSII participants, a longer history of rotating night shift work was suggestively associated with an elevated risk for incident CVD and was consistently associated with higher all-cause mortality risk. The robustness of these findings was supported by detailed covariate adjustment, evaluation of dose-response relationships, and use of multiple alternative shift work metrics in secondary analyses, and in line with our earlier findings in this cohort, which were based on a shorter follow-up period (5, 6). Similarly, greater eveningness was modestly associated with higher risks for both CVD risk and all-cause mortality. Although we did not observe statistically significant differences in the association between rotating night shift work duration and CVD risk or all-cause mortality across chronotype groups, the overall patterns of the associations as shown by both the categorical analysis and the restricted cubic spline analyses were consistent with our hypothesis that individuals with a morning chronotype appeared more vulnerable to the adverse health impact by rotating night shift work.

### Interpretation

Evidence linking shift work with incident CVD and all-cause mortality is relatively well established. Multiple systematic reviews and meta-analyses have reported increased risks of vascular events and mortality associated with shift work (44–46), and one review further suggested that the excess risk of CVD becomes more apparent after the first five years of shift work (44). In contrast, the association between chronotype and these outcomes has been less extensively studied. A broad umbrella review of sleep traits and a chronotype-specific systematic review both suggested that greater eveningness is associated with a less favorable cardiometabolic profile, with less conclusive evidence for CVD incidence (47, 48). Consistent with this literature, our previous studies in NHSII reported adverse associations of greater eveningness with cardiovascular risk factors, such as lower diet quality and abnormal blood lipids, as well as higher risks of incident diabetes and CVD (18, 19, 49, 50).

Fewer studies have examined the interaction between shift work and chronotype in relation to health outcomes. Motivated by experimental evidence supporting adverse metabolic and cardiovascular consequences of circadian misalignment (13), and evidence from the NHSII cohort, suggesting that poor alignment between rotating shift work and chronotype may lead to greater disruption of melatonin rhythms (51), our study suggests stronger associations of rotating night shift work with incident CVD and all-cause mortality among definite morning types, broadly consistent with the hypothesis that work schedule-chronotype mismatch may exacerbate the adverse consequences associated with rotating night shift work. However, the patterns differed for each outcome: for incident CVD, a stronger linear trend in risk with increasing duration of rotating night shift work was observed among morning chronotypes, whereas for all-cause mortality, morning chronotypes showed an elevated risk at shorter durations of shift work that appeared to attenuate at longer durations. Because NHSII better captures the mismatch between morning chronotype and rotating night work than between evening chronotype and early morning schedules, this pattern may also have been more readily detectable in our data.

In a systematic review of 16 studies examining chronotype in relation to shift-work tolerance, nine studies reported better tolerance among individuals with greater eveningness, although these studies primarily focused on outcomes such as sleepiness, sleep disturbance, fatigue, and related psychosocial problems, and sample sizes were small (52). This may potentially explain the observed plateau in all-cause mortality among morning chronotypes, as individuals with shorter shift work durations may have been less tolerant of night shift work and therefore more likely to have been discontinued such work schedules earlier, possibly due to emerging health issues or a greater propensity to develop shift work disorder (53).

While work schedule-chronotype mismatch has been consistently associated with shorter sleep duration, greater social jet lag, and other sleep disturbances (21, 54), evidence for its associations with cardiometabolic risk factors has been more mixed. Notably, some cross-sectional studies reported stronger associations between shift work and cardiovascular risk factors among evening chronotypes (23, 55), whereas findings from larger prospective studies have been more consistent with our results. In a prior NHSII study, the association between rotating night shift work and incident type 2 diabetes differed by chronotype and duration of shift work, with risk increasing with longer shift work exposure among definite morning chronotypes (20). Likewise, a prospective study of more than 36,000 UK Biobank participants with hypertension found stronger associations between night shift work and cardiometabolic multimorbidity among morning chronotypes (56). Other large prospective UK Biobank studies generally found larger effect estimates associated with shift work for cardiovascular-kidney-metabolic disease, CVD, or all-cause mortality among morning chronotypes, although most studies did not focus on the mismatch or formally examine the subgroup differences by chronotype (57–59). Taken together, the prospective literature on cardiometabolic and mortality outcomes appears mixed but tends to align more closely with our findings, with limited evidence of formal interaction overall but some suggestion that morning chronotypes may be more susceptible to night shift work.

A plausible mechanism linking work schedule-chronotype mismatch with incident CVD and all-cause mortality is greater circadian misalignment, whereby sleep, light exposure, food intake, and physical activity occur at biologically unfavorable times (60). Prior reviews have highlighted that circadian disruption can adversely affect glucose metabolism, blood pressure, autonomic and vascular function, inflammation, and molecular clock pathways relevant to cardiovascular disease, providing a biologically plausible basis for these associations (9, 10, 15, 61, 62). Supporting a chronotype-specific pathway, a sub-study of 130 NHSII nurses found that associations of rotating night shift work with urinary melatonin rhythm parameters differed by chronotype, suggesting less disrupted melatonin rhythms under better chronotype-schedule alignment (51). Similarly, in the Maastricht Study, alignment of weekday meal timing and physical activity with chronotype was associated with more favorable glucose metabolism (63). An intervention study further reported that chronotype-adjusted shift schedules improved sleep in shift workers, suggesting that better alignment between chronotype and work schedule may help mitigate pathways relevant to longer-term cardiometabolic risk (54).

Despite the lack of significant statistical interactions, our results suggest a more complex pattern in the associations of rotating night shift work with CVD and all-cause mortality across chronotype categories, with morning chronotypes potentially more vulnerable; specifically, the association was more linear for incident CVD, whereas all-cause mortality showed a sharper nonlinear increase followed by a plateau. Recent Mendelian randomization evidence suggests bidirectional genetic associations between chronotype and shift work, while other studies have reported more favorable sleep duration among evening chronotypes who consistently work night shifts and contributions of both phenotypic and genotypic characteristics to adaptation to shift work (64–66). These studies also raise the possibility that reverse causation and healthy worker bias may have attenuated the observed subgroup differences by chronotype. Individuals who are less tolerant of rotating night shift work may be more likely to leave such schedules over time, resulting in a selected group of workers who are, on average, more tolerant of this exposure and potentially at lower risk for adverse health outcomes.

### Strengths

Strengths of our study include the large population-based sample, prospective cohort design, long-term follow-up, and high retention in NHSII. The biennial questionnaires provided repeated assessment, which captured the time-varying nature of shift work and other key variables, potentially reducing measurement errors and residual confounding. In addition, many questionnaire-based measures in NHSII, including lifestyle and health factors, have been extensively validated in prior work (37, 39, 67). Ascertainment of incident medical conditions in NHSII is supported by rigorous follow-up procedures, including supplementary questionnaires and confirmation using medical record review when applicable, which strengthens outcome validity. Finally, NHSII is part of the Nurses’ Health Studies, which have been widely recognized as a major platform for population health discovery and translation to prevention and public health guidance (68). As an occupational cohort with substantial exposure to rotating night shift work, it also provides a valuable setting to study work schedule-chronotype mismatch.

## Limitations

Caution should be exercised when generalizing our findings. NHSII participants are primarily white female registered nurses with relatively high education and socioeconomic status in a similar occupational setting; therefore, results may not apply to other segments of the population. In addition, shift scheduling practices and workplace conditions may have changed over time, so generational differences may limit applicability to current shift-work environments.

Other limitations include potential exposure misclassification and incomplete characterization of circadian misalignment. We focused on rotating night shift work and did not capture other work schedules that may also disrupt circadian timing, including early morning shifts; thus, the reference group may have included participants exposed to other forms of shift work, and we could not directly evaluate mismatch between evening chronotype and early morning work. Rotating night shift work was self-reported using categorical responses, which may introduce measurement error and recall bias; however, shift work was assessed repeatedly using standardized NHSII questions, and we tested alternative exposure definitions to assess robustness. Chronotype reflects circadian preference but may not reliably capture the underlying circadian phase, which is more biologically relevant to shift work misalignment. It was also assessed with a single item, which may introduce measurement error; however, this measure has shown validity against dim light melatonin onset in prior studies (69, 70), and we incorporated repeated chronotype assessment. Developing scalable metrics that more reliably capture circadian phase and misalignment is essential for future studies.

Additionally, selection bias remains possible because the analytic sample required participation in 2009 with self-reported chronotype, although overall follow-up in NHSII is high (25). A healthy worker survivor effect, whereby participants in poorer health are more likely to leave rotating night work, is also plausible; we partially addressed this by examining current versus past shift work and time since quitting. Moreover, although we adjusted for a wide range of time-varying covariates, residual confounding by unmeasured factors is still possible. Finally, although the cohort was large, the number of incident CVD events was modest for sub-group analysis, which may have limited statistical power to detect significant interaction despite patterns that were broadly consistent with our hypothesis (71).

## Conclusion

Taken together, in this prospective analysis of NHSII participants, a longer history of rotating night shift work was consistently associated with higher all-cause mortality, with evidence of a dose-response pattern, and was suggestively associated with an elevated risk for incident CVD. Greater eveningness was also modestly associated with higher risks for both incident CVD and all-cause mortality. Although we did not observe statistically significant evidence that chronotype modified the associations of rotating night shift work with incident CVD or all-cause mortality, the overall patterns from both categorical and restricted cubic spline analyses were consistent with our hypothesis that individuals with a morning chronotype may be more vulnerable to the adverse health impacts of rotating night shift work. From a public health and clinical perspective, these findings support efforts to reduce chronic exposure to rotating night shift work where feasible and to take into account chronotype in shift work scheduling for risk assessment and health promotion.

## Acknowledgements

The Nurses’ Health Study II has been supported by the National Cancer Institute and the National Heart, Lung, and Blood Institute of the National Institutes of Health under award numbers U01CA176726, U01HL145386, and R01HL088521. This work was additionally supported by the National Heart, Lung, and Blood Institute under award number R01HL155395. S.K. was supported by American Heart Association grant 24POST1188091 (2024 Postdoctoral Fellowship). F.A.J.L.S. was supported by National Heart, Lung, and Blood Institute grants R01HL153969 and R01HL164454. T.H. was supported by the Intramural Research Program of the National Institute on Aging, National Institutes of Health (ZIAAG000530). E.S. was supported by the European Research Council under the European Union’s Horizon Europe Research and Innovation Programme (ERC Advanced Grant CLOCKrisk, Grant Agreement No. 101053225). The content is solely the responsibility of the authors and does not necessarily represent the official views of the National Institutes of Health. Views and opinions expressed are, however, those of the authors only and do not necessarily reflect those of the European Union or the European Research Council Executive Agency. Neither the European Union nor the granting authority can be held responsible for them.

The authors would like to acknowledge the contribution to this study from central cancer registries supported through the Centers for Disease Control and Prevention’s National Program of Cancer Registries (NPCR) and/or the National Cancer Institute’s Surveillance, Epidemiology, and End Results (SEER) Program. Central registries may also be supported by state agencies, universities, and cancer centers. Participating central cancer registries include the following: Alabama, Alaska, Arizona, Arkansas, California, Delaware, Colorado, Connecticut, Florida, Georgia, Hawaii, Idaho, Indiana, Iowa, Kentucky, Louisiana, Maine, Maryland, Massachusetts, Michigan, Mississippi, Montana, Nebraska, Nevada, New Hampshire, New Jersey, New Mexico, New York, North Carolina, North Dakota, Ohio, Oklahoma, Oregon, Pennsylvania, Puerto Rico, Rhode Island, Seattle SEER Registry, South Carolina, Tennessee, Texas, Utah, Virginia, West Virginia, Wyoming.

## Previous Presentation

A preliminary version of this work was presented at the 57^th^ Annual Meeting of the Society for Epidemiologic Research; June 18-21, 2024; Austin, Texas.

## Data Availability Statement

Data from the Nurses’ Health Study II are not publicly available because of participant confidentiality and consent restrictions. Information regarding data access and collaboration is available through the Nurses’ Health Studies website. Investigators interested in accessing the data may submit a research proposal according to established Nurses’ Health Studies procedures.

---

**Supplemental Table 1.** Prospective associations of alternative rotating night shift work exposures with risk of cardiovascular disease, N = 69,458.

|  |  |  |  |  |  |
| --- | --- | --- | --- | --- | --- |
|  | Cases / Person-years | Crude incidence per 100,000 PY | Hazard Ratio (95% CI) |  |  |
|  |  |  | Age-adjusted model | Model 1* | Model 2† |
| Duration of rotating night shift work (4 categories) |  |  |  |  |  |
| - None | 265 / 217194 | 122 | Ref. | Ref. | Ref. |
| - <5 years | 459 / 394657 | 116 | 1.00 (0.86, 1.16) | 0.98 (0.84, 1.14) | 0.95 (0.81, 1.10) |
| - ≥5 to <10 years | 166 / 114023 | 146 | **1.31 (1.08, 1.60)** | **1.24 (1.02, 1.51)** | 1.11 (0.91, 1.35) |
| - ≥10 years | 115 / 74757 | 154 | **1.26 (1.01, 1.57)** | 1.16 (0.93, 1.46) | 1.00 (0.80, 1.26) |
| Duration of rotating night shift work (complete case only: n = 22,794) ‡ |  |  |  |  |  |
| - None | 89 / 75960 | 117 | Ref. | Ref. | Ref. |
| - <5 years | 137 / 132115 | 104 | 0.91 (0.70, 1.19) | 0.90 (0.69, 1.18) | 0.85 (0.65, 1.11) |
| - ≥5 years | 83 / 57732 | 144 | 1.33 (0.98, 1.80) | 1.26 (0.93, 1.71) | 1.09 (0.80, 1.49) |
| Per 5-year increase | 309 / 265807 | 116 | **1.13 (1.00, 1.27)** | 1.11 (0.98, 1.25) | 1.05 (0.93, 1.19) |
| P Value for Trend | NA | NA | **0.0423** | 0.1008 | 0.4114 |
| Ever shift work |  |  |  |  |  |
| - Yes | 740 / 583437 | 127 | 1.09 (0.95, 1.26) | 1.05 (0.91, 1.21) | 0.98 (0.85, 1.14) |
| - No | 265 / 217194 | 122 | Ref. | Ref. | Ref. |
| Time since quitting rotating night shift work § |  |  |  |  |  |
| - Current | 77 / 51332 | 150 | Ref. | Ref. | Ref. |
| - <12 years | 159 / 112907 | 141 | 0.93 (0.71, 1.22) | 0.95 (0.72, 1.25) | 0.96 (0.73, 1.27) |
| - 12 to 24 years | 209 / 167404 | 125 | 0.83 (0.64, 1.08) | 0.87 (0.66, 1.15) | 0.92 (0.69, 1.21) |
| - >24 years | 295 / 251795 | 117 | **0.68 (0.53, 0.88)** | 0.77 (0.58, 1.02) | 0.88 (0.66, 1.17) |
| Per 5-year increase § | 740 / 583437 | 127 | **0.95 (0.92, 0.97)** | **0.96 (0.93, 0.99)** | 0.98 (0.95, 1.01) |
| P Value for Trend § | NA | NA | <.0001 | 0.0096 | 0.1911 |
| Shift work status |  |  |  |  |  |
| - Never shift worked | 265 / 217194 | 122 | Ref. | Ref. | Ref. |
| - Current | 77 / 51332 | 150 | **1.40 (1.08, 1.80)** | 1.30 (1.00, 1.70) | 1.12 (0.86, 1.46) |
| - Past shift work | 663 / 532105 | 125 | 1.07 (0.92, 1.23) | 1.03 (0.90, 1.19) | 0.97 (0.84, 1.12) |
| Night shift work frequency in 2009 (n = 65,278) |  |  |  |  |  |
| - None | 772 / 650522 | 119 | Ref. | Ref. | Ref. |
| - 1-6 nights per month | 84 / 54824 | 153 | **1.38 (1.10, 1.73)** | **1.31 (1.04, 1.65)** | 1.25 (0.99, 1.57) |
| - ≥7 nights/month | 67 / 47943 | 140 | 1.20 (0.94, 1.55) | 1.15 (0.88, 1.48) | 0.97 (0.75, 1.26) |
| Per 5 nights/month increase | 923 / 753289 | 123 | 1.07 (0.99, 1.16) | 1.05 (0.97, 1.14) | 0.99 (0.91, 1.08) |
| P Value for Trend | NA | NA | 0.0843 | 0.2628 | 0.7991 |
| * Model 1: Age, ethnic background, census-tract median family income, marital status, workplace, family history of MI and/or stroke, menopausal status, and postmenopausal hormone therapy. |  |  |  |  |  |
| † Model 2: Model 1 + history of diabetes/hypertension/hypercholesterolemia, aspirin use, smoking status, sleep duration, physical activity, body mass index (BMI), Alternate Healthy Eating Index-2010 score (AHEI-2010), and alcohol use. Census-tract median family income, AHEI-2010 score, BMI, physical activity, and alcohol use were modeled as continuous variables. |  |  |  |  |  |
| ‡ This shift work variable was assessed through the 2007 questionnaire cycle and treated as a fixed (non–time-dependent) variable. Participants with missing data on shift work duration in 2007 or any earlier cycle were excluded from the analysis. All other shift work variables were time-dependent, assessed at baseline in 2009 and subsequently updated in 2011 and 2013. |  |  |  |  |  |
| § Among current and former shift workers (n = 50,737). NA: Not Applicable |  |  |  |  |  |

**Supplemental Table 2.** Prospective associations of alternative rotating night shift work exposures with all-cause mortality, N = 69,458.

|  |  |  |  |  |  |
| --- | --- | --- | --- | --- | --- |
|  | Cases /<br>Person-years | Crude incidence<br>per 100,000 PY | Hazard Ratio (95% CI) |  |  |
|  |  |  | Age-adjusted<br>model | Model 1* | Model 2† |
| **Duration of rotating night shift work (4 categories)** |  |  |  |  |  |
| - None | 406 / 218577 | 186 | Ref. | Ref. | Ref. |
| - <5 years | 863 / 397144 | 217 | **1.23 (1.09, 1.38)** | **1.20 (1.07, 1.36)** | **1.17 (1.04, 1.32)** |
| - 5-10 years | 277 / 114979 | 241 | **1.44 (1.23, 1.67)** | **1.36 (1.17, 1.59)** | **1.29 (1.10, 1.50)** |
| - ≥10 years | 231 / 75343 | 307 | **1.66 (1.41, 1.95)** | **1.51 (1.28, 1.77)** | **1.40 (1.18, 1.65)** |
| **Duration of rotating night shift work (complete case only: n = 22,794) ‡** |  |  |  |  |  |
| - None | 122 / 76424 | 160 | Ref. | Ref. | Ref. |
| - <5 years | 247 / 132821 | 186 | 1.21 (0.97, 1.50) | 1.19 (0.95, 1.48) | 1.17 (0.94, 1.46) |
| - ≥5 years | 132 / 58206 | 227 | **1.48 (1.15, 1.91)** | **1.38 (1.07, 1.78)** | **1.32 (1.03, 1.71)** |
| Per 5-year increase | 501 / 267452 | 187 | **1.20 (1.11, 1.31)** | **1.18 (1.08, 1.28)** | **1.17 (1.07, 1.27)** |
| P Value for Trend | NA | NA | **<.0001** | **0.0003** | **0.0007** |
| **Ever shift work** |  |  |  |  |  |
| - Yes | 1371 / 587466 | 233 | **1.32 (1.18, 1.48)** | **1.27 (1.14, 1.43)** | **1.22 (1.09, 1.37)** |
| - No | 406 / 218577 | 186 | Ref. | Ref. | Ref. |
| **Time since quitting rotating night shift work §** |  |  |  |  |  |
| - Current | 104 / 51732 | 201 | Ref. | Ref. | Ref. |
| - <12 years | 290 / 113677 | 255 | 1.16 (0.92, 1.45) | 1.20 (0.95, 1.51) | 1.19 (0.95, 1.50) |
| - 12 to 24 years | 393 / 168625 | 233 | 1.01 (0.81, 1.25) | 1.08 (0.86, 1.36) | 1.10 (0.87, 1.38) |
| - >24 years | 584 / 253433 | 230 | 0.90 (0.73, 1.11) | 1.03 (0.82, 1.30) | 1.11 (0.88, 1.40) |
| Per 5-year increase § | 1371 / 587466 | 233 | **0.97 (0.95, 0.99)** | 0.99 (0.97, 1.01) | 1.00 (0.98, 1.02) |
| P Value for Trend § | NA | NA | **0.0016** | 0.2837 | 0.9720 |
| **Shift work status** |  |  |  |  |  |
| - Never shift worked | 406 / 218577 | 186 | Ref. | Ref. | Ref. |
| - Current | 104 / 51732 | 201 | **1.36 (1.09, 1.69)** | **1.31 (1.05, 1.64)** | 1.21 (0.97, 1.51) |
| - Past shift work | 1267 / 535735 | 237 | **1.32 (1.18, 1.48)** | **1.27 (1.14, 1.42)** | **1.22 (1.09, 1.37)** |
| **Night shift work frequency in 2009 (n = 65,278)** |  |  |  |  |  |
| - None | 1343 / 654714 | 205 | Ref. | Ref. | Ref. |
| - 1-6 nights per month | 111 / 55250 | 201 | 1.02 (0.84, 1.23) | 1.02 (0.84, 1.24) | 1.00 (0.82, 1.21) |
| - ≥7 nights/month | 135 / 48301 | 280 | **1.39 (1.17, 1.66)** | **1.34 (1.12, 1.61)** | **1.22 (1.02, 1.47)** |
| Per 5 nights/month increase | 1589 / 758265 | 210 | **1.12 (1.06, 1.19)** | **1.10 (1.04, 1.17)** | **1.06 (1.00, 1.13)** |
| P Value for Trend | NA | NA | **0.0001** | **0.0016** | **0.0465** |
| * Model 1: Age, ethnic background, census-tract median family income, marital status, workplace, family history of MI and/or stroke, menopausal status, and postmenopausal hormone therapy. |  |  |  |  |  |
| † Model 2: Model 1 + history of diabetes/hypertension/hypercholesterolemia, aspirin use, smoking status, sleep duration, physical activity, body mass index (BMI), Alternate Healthy Eating Index-2010 score (AHEI-2010), and alcohol use. Census-tract median family income, AHEI-2010 score, BMI, physical activity, and alcohol use were modeled as continuous variables. |  |  |  |  |  |
| ‡ This shift work variable was assessed through the 2007 questionnaire cycle and treated as a fixed (non–time-dependent) variable. Participants with missing data on shift work duration in 2007 or any earlier cycle were excluded from the analysis. All other shift work variables were time-dependent, assessed at baseline in 2009 and subsequently updated in 2011 and 2013. |  |  |  |  |  |
| § Among current and former shift workers (n = 50,738). NA: Not Applicable |  |  |  |  |  |

**Supplemental Table 3.** Prospective association between chronotype, rotating night shift work, and myocardial infarction, NHSII (N = 69,458)

|  |  |  |  |  |  |
| --- | --- | --- | --- | --- | --- |
|  |  |  | Hazard Ratio (95% CI) |  |  |
|  | Cases /<br>Person-years | Crude incidence<br>per 100,000 PY | Age-adjusted<br>model | Model 1* | Model 2† |
| **Duration of rotating night shift work** |  |  |  |  |  |
| - None | 170 / 217317 | 78 | Ref. | Ref. | Ref. |
| - <5 years | 300 / 394840 | 76 | 1.02 (0.84, 1.23) | 0.99 (0.82, 1.20) | 0.95 (0.78, 1.15) |
| - ≥5 years | 184 / 188895 | 97 | **1.32 (1.07, 1.63)** | 1.22 (0.98, 1.51) | 1.05 (0.85, 1.30) |
| Per 5-year increase | 654 / 801052 | 82 | **1.11 (1.03, 1.19)** | 1.07 (1.00, 1.16) | 1.02 (0.94, 1.10) |
| P Value for Trend | NA | NA | **0.0060** | 0.0647 | 0.6369 |
| **Chronotype** |  |  |  |  |  |
| - Definite morning | 207 / 268866 | 77 | 0.95 (0.80, 1.13) | 0.97 (0.81, 1.15) | 1.06 (0.89, 1.26) |
| - Intermediate (neither and more) | 340 / 435257 | 78 | Ref. | Ref. | Ref. |
| - Definite evening | 107 / 96928 | 110 | **1.40 (1.12, 1.74)** | **1.33 (1.07, 1.65)** | 1.11 (0.89, 1.39) |
| Per 1-level increase toward evening | 654 / 801052 | 82 | **1.18 (1.05, 1.33)** | **1.14 (1.02, 1.29)** | 1.01 (0.90, 1.14) |
| P Value for Trend | NA | NA | **0.0061** | **0.0258** | 0.8850 |
| * Model 1: Age, ethnic background, census-tract median family income, marital status, workplace, family history of MI and/or stroke, menopausal status, and postmenopausal hormone therapy. |  |  |  |  |  |
| † Model 2: Model 1 + history of diabetes/hypertension/hypercholesterolemia, aspirin use, smoking status, sleep duration, physical activity, body mass index (BMI), Alternate Healthy Eating Index-2010 score (AHEI-2010), and alcohol use. Census-tract median family income, AHEI-2010 score, BMI, physical activity, and alcohol use were modeled as continuous variables. |  |  |  |  |  |
| NA: Not Applicable |  |  |  |  |  |

**Supplemental Table 4.**
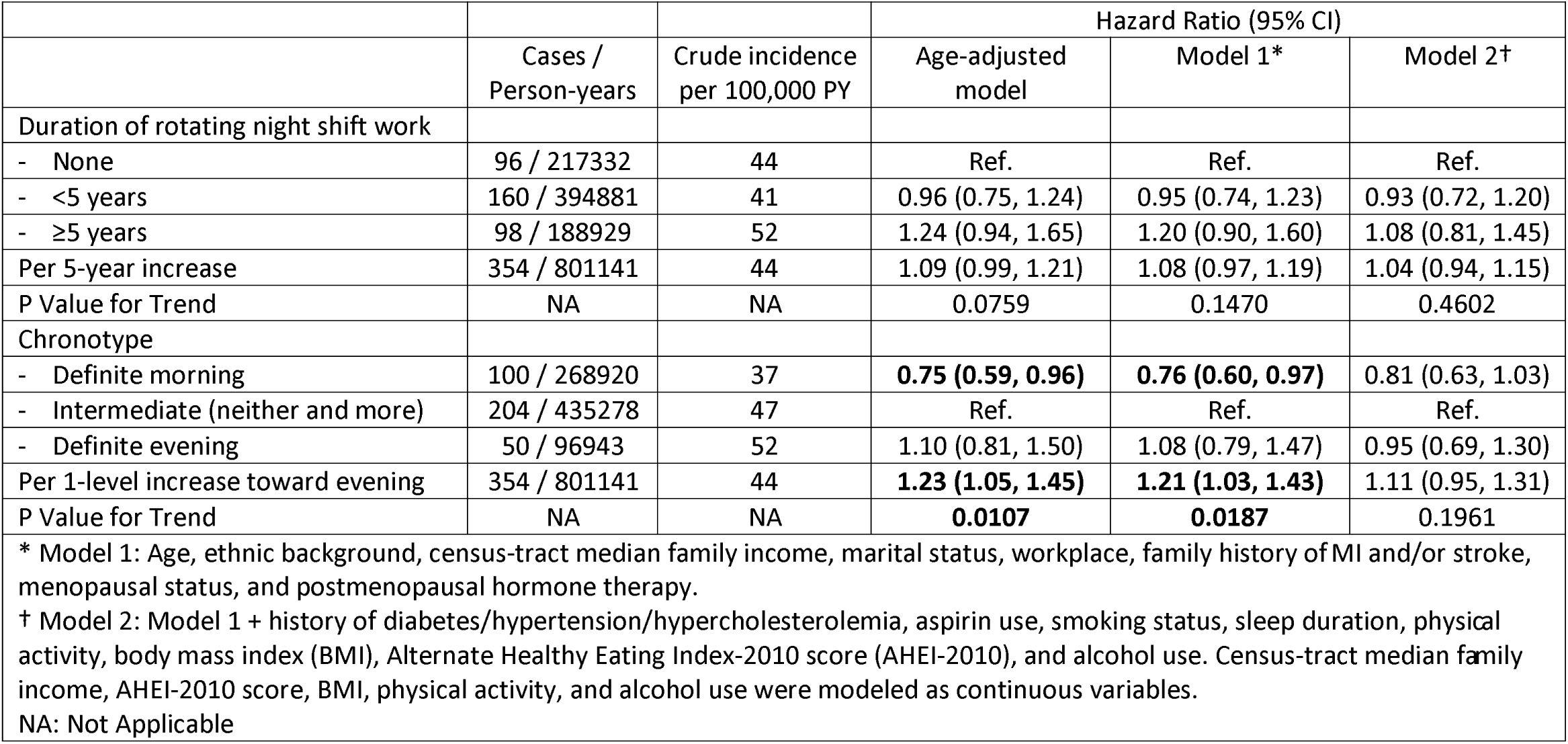
Prospective association between chronotype, rotating night shift work, and stroke, NHSII (N = 69,458)

**Supplemental Table 5.** Joint association of shift work and chronotype with risks of cardiovascular disease and all-cause mortality in the NHSII (N = 69,458)

|  | Duration of rotating night shift work |  |  |  |  |  |
| --- | --- | --- | --- | --- | --- | --- |
|  | None |  | <5 years |  | ≥5 years |  |
|  | Association with Incident CVD |  |  |  |  |  |
| Chronotype | Cases / PY;<br>Crude incidence | HR<br>(95% CI) | Cases / PY;<br>Crude incidence | HR<br>(95% CI) | Cases / PY;<br>Crude incidence | HR<br>(95% CI) |
| Definite morning | 90 / 76394; 118 | 0.95 (0.73, 1.23) | 133 / 133720; 99 | 0.82 (0.64, 1.03) | 84 / 58634; 143 | 1.18 (0.90, 1.55) |
| Intermediate | 143 / 118685; 120 | Reference | 247 / 214447; 115 | 0.99 (0.81, 1.22) | 151 / 101882; 148 | 1.20 (0.95, 1.52) |
| Definite evening | 32 / 22115; 145 | 1.15 (0.79, 1.70) | 79 / 46491; 170 | **1.38 (1.05, 1.82)** | 46 / 28263; 163 | 1.29 (0.92, 1.80) |
|  | Association with All-Cause Mortality |  |  |  |  |  |
| Definite morning | 116 / 76841; 151 | **0.75 (0.60, 0.94)** | 272 / 134380; 202 | 1.03 (0.86, 1.22) | 128 / 59100; 217 | 1.08 (0.87, 1.34) |
| Intermediate | 240 / 119454; 201 | Reference | 457 / 215874; 212 | 1.08 (0.93, 1.27) | 273 / 102641; 266 | **1.30 (1.09, 1.55)** |
| Definite evening | 50 / 22283; 224 | 0.97 (0.71, 1.31) | 134 / 46890; 286 | **1.32 (1.07, 1.64)** | 107 / 28581; 374 | **1.69 (1.34, 2.14)** |

Models adjusted for age, ethnic background, census-tract median family income, marital status, workplace, family history of MI and/or stroke, menopausal status, and postmenopausal hormone therapy.
HR: Hazard Ratio; PY: Person-Year

**Supplemental Figure 1.**
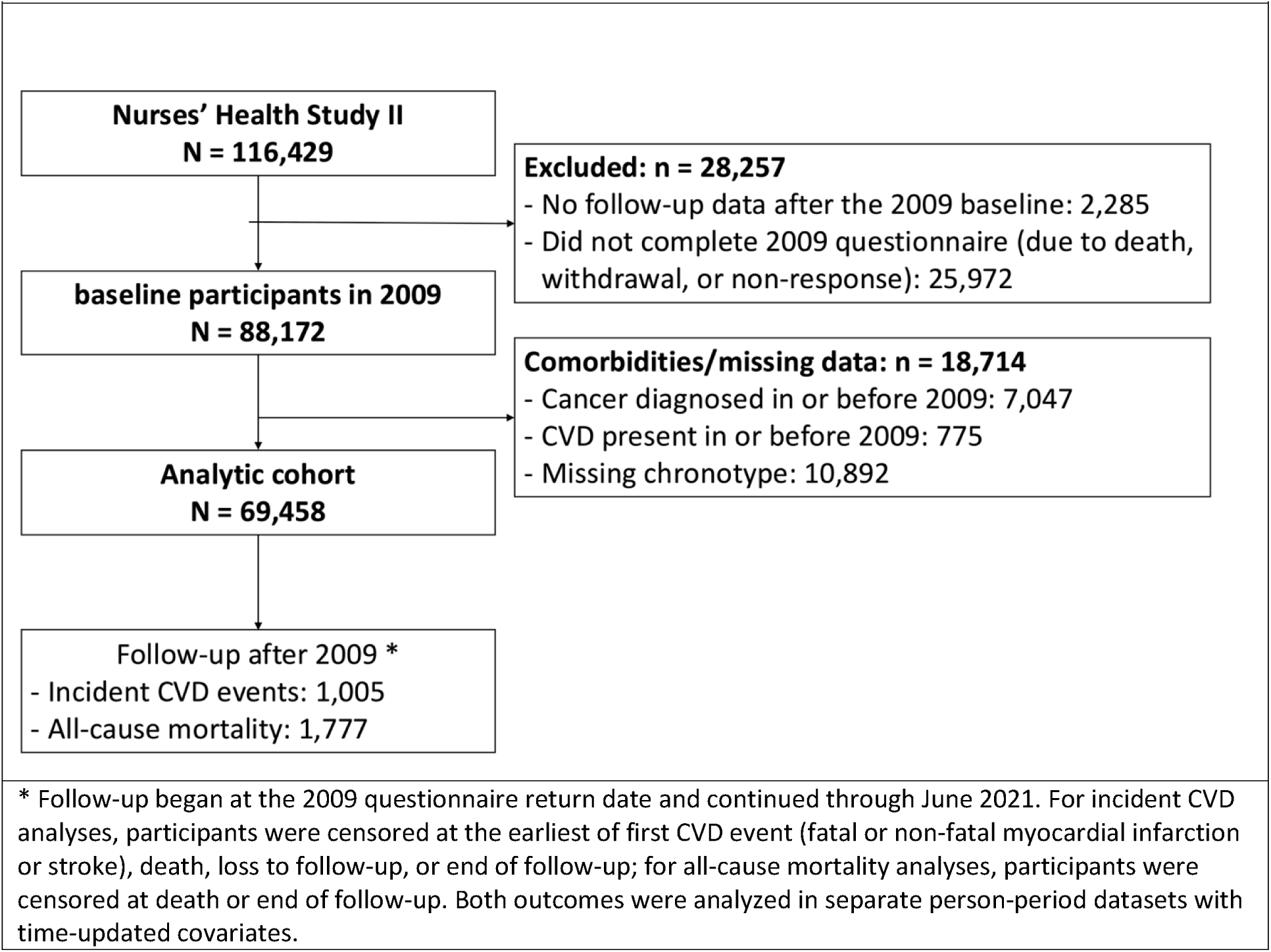
Study flow diagram.

**Supplemental Figure 2.**
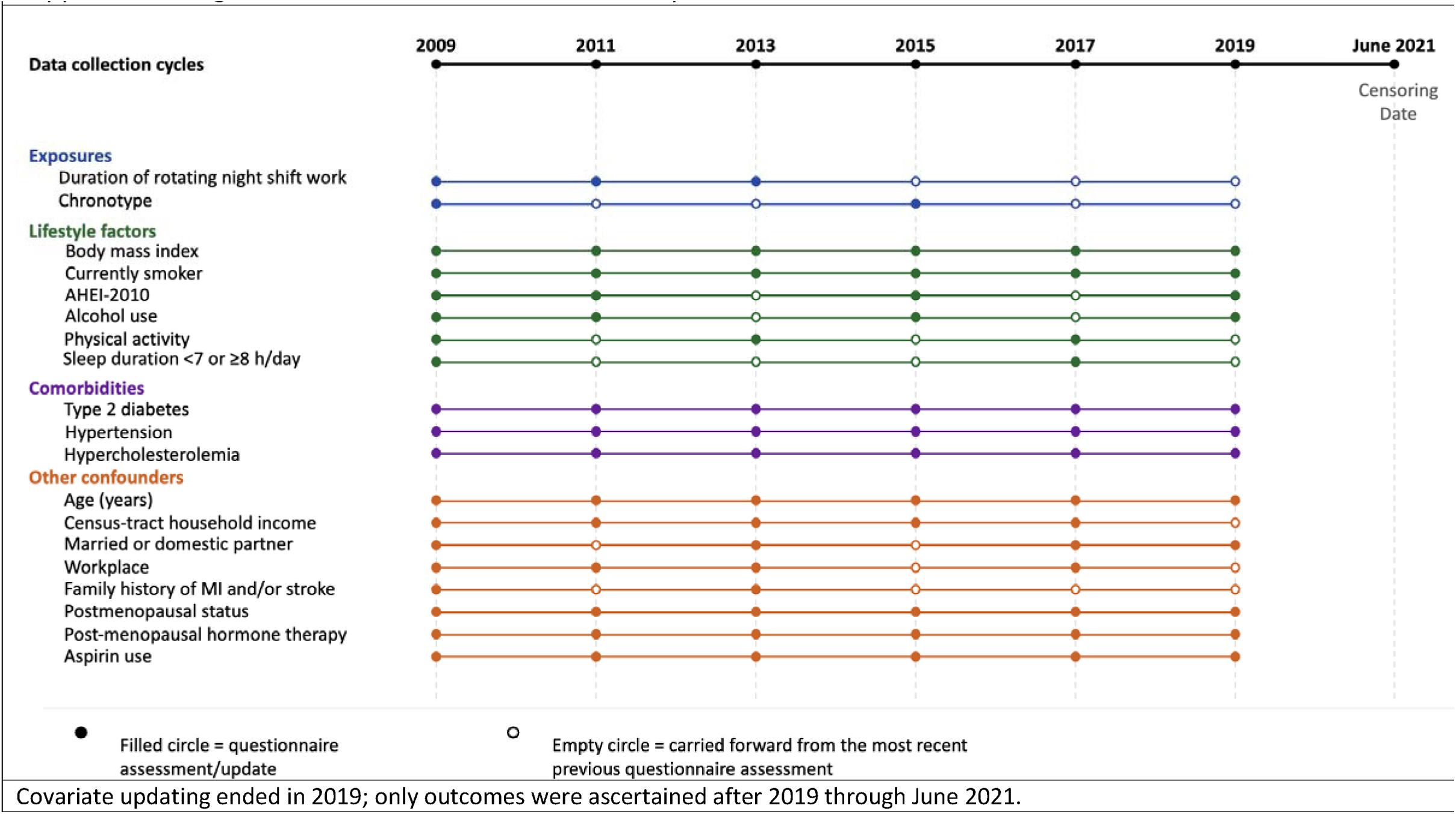
Data collection timeline for exposures, covariates, and outcome ascertainment.

